# Response to COVID-19 booster vaccinations in seronegative people with MS

**DOI:** 10.1101/2022.03.12.22272083

**Authors:** Emma C Tallantyre, Martin J Scurr, Nicola Vickaryous, Aidan Richards, Valerie Anderson, David Baker, Randy Chance, Nikos Evangelou, Katila George, Gavin Giovannoni, Katharine E Harding, Aimee Hibbert, Gillian Ingram, Stephen Jolles, Meleri Jones, Angray S Kang, Samantha Loveless, Stuart J Moat, Neil P Robertson, Francesca Rios, Klaus Schmierer, Mark Willis, Andrew Godkin, Ruth Dobson

**Author notes:** Corresponding author: Dr Ruth Dobson, Preventive Neurology Unit, Wolfson Institute of Population Health, Charterhouse Square, EC1M 6BQ, @drruthdobson. These authors contributed equally (Martin J Scurr, Nicola Vickaryous, Aidan Richards).

## Abstract

**Background:** Uncertainties remain about the benefit of a 3rd COVID-19 vaccine for people with attenuated response to earlier vaccines. This is of particular relevance for people with multiple sclerosis (pwMS) treated with anti-CD20 therapies and fingolimod, who have substantially reduced antibody responses to initial vaccine course.

**Methods:** PwMS taking part in a seroprevalence study without a detectable IgG response following COVID-19 vaccines 1&2 were invited to participate. Participants provided a dried blood spot +/-venous blood sample 2-12 weeks following COVID-19 vaccine 3. Humoral and T cell responses to SARS-CoV-2 spike protein and nucleocapsid antigen were measured. The relationship between evidence of prior COVID-19 infection and immune response to COVID-19 vaccine 3 was evaluated using Fishers exact test.

**Results:** Of 81 participants, 79 provided a dried blood spot sample, of whom 38 also provided a whole blood sample; 2 provided only whole blood. Anti-SARS-CoV-2-spike IgG seroconversion post-COVID-19 vaccine 3 occurred in 26/79 (33%) participants; 26/40 (65%) had positive T-cell responses. Overall, 31/40 (78%) demonstrated either humoral or cellular immune response post-COVID-19 vaccine 3. There no association between laboratory evidence of prior COVID-19 infection and anti-spike seroconversion following COVID-19 vaccine 3.

**Conclusions:** Approximately one third of pwMS who were seronegative after initial COVID-19 vaccination seroconverted after booster (third) vaccination, supporting the use of boosters in this group. Almost 8 out of 10 had a measurable immune response following 3rd COVID-19 vaccine.

**Key messages:** *What is already known:* The benefits of COVID vaccination are well described. It is unknown whether there is additional benefit afforded from a third COVID-19 vaccination in those people who have failed to mount a serological response to their initial vaccine course.

*What this study adds:* Approximately one third of people with MS in our study, all of whom had failed to response to initial vaccine course, developed anti-spike antibodies following a third COVID-19 vaccine. Two-thirds of participants had T cell response to vaccination. No people taking fingolimod appeared to mount a T cell response to vaccination.

*How this study might influence practice:* These findings highlight potential benefits of booster vaccinations to a substantial proportion of immunosuppressed people who have failed to respond to initial vaccination course. The clinical correlates of antibody and T-cell responses to COVID-19 remain uncertain but they are almost certainly associated with milder subsequent disease in the general population.

## Introduction

To date, the COVID-19 pandemic has resulted in over 5 million deaths worldwide[1]. Vaccines have reduced overall COVID-19 associated morbidity and mortality, but uncertainties remain about the benefit of COVID-19 vaccination for people with primary and secondary immunocompromise. We and others have demonstrated that many people with multiple sclerosis (pwMS) treated with anti-CD20 and Sphingosine-1-phosphate (S1P) modulators have an attenuated immune response to the first two doses of COVID-19 vaccination[2][3]. Studies have so far largely focused on humoral immune responses. However, the impact of booster vaccination in pwMS who mount an inadequate response to initial COVID-19 vaccination is not known. We set out to report humoral and T-cell responses following a third COVID-19 vaccination in pwMS who were known to be seronegative after their second COVID-19 vaccine.

## Methods

A subgroup of pwMS taking part in a seroprevalence study[2], were invited to participate. Selection criteria were IgG negative anti-spike COVID-19 antibody status at week 4-8 post-COVID-19 vaccine 2, and willingness to provide a further blood sample 2-12 weeks following COVID-19 vaccine 3. All participants were invited to provide a dried blood spot and a whole blood sample. Dried blood spot samples were collected as previously described [2] during November 2021-January 2022. Where participants were willing to attend for blood sampling, a whole blood sample was collected on the same day. Data on demographics, MS type and treatment, and COVID-19 infection/vaccine dates were derived from the medical notes during January-February 2022. All participants provided written informed consent to take part in this study. This study has Research Ethics Committee approval (REC 19/WA/0058 (Wales REC 3 – covering samples processed in Cardiff), 05/WSE03/111 (South East Wales REC – covering samples processed in Cardiff) and 20/NE/0176 (Newcastle North Tyneside REC – covering QMUL samples).

Humoral responses to SARS-CoV-2 (S1 subunit of the spike protein) following COVID-19 vaccine 3 were measured on dried blood spots using the FDA-approved EuroImmun (PerkinElmer) enzyme-linked immunosorbent assay (ELISA) as per manufacturer instructions. Validation of the EuroImmun assay demonstrates that plasma/serum and DBS specimens produce equivalent results [4]. Results are expressed as a ratio of the optical density (OD) of the participant sample over the OD of the calibrator: ratio <0.8 negative, </=0.8 to <1.1 borderline, >/=1.1 positive. This assay provided good agreement with the assays used in our previous study (Kantaro vs EuroImmun, n=23, R^2^=0.94, kappa=1.0; Globody vs EuroImmun, n=23, R^2^=0.81, kappa=0.83).

In the subset of 40 participants who provided whole blood samples, humoral responses were also measured using the Bio-Plex Pro Human SARS-CoV-2 (N/RBD/S1/S2 subunits of spike protein) alongside the VIROTROL SARS-CoV-2 single-level control (Bio-Rad) and performed according to manufacturer’s instructions on a Bio-Plex 200 (Bio-Rad). T-cell responses to SARS-CoV-2 were measured in these participants, using a commercially available whole blood assay (ImmunoServ Ltd), as previously described[4]. Briefly, 10ml heparinised venous blood from each patient was collected and processed within 24 hours of blood draw. Whole blood samples were stimulated with peptides (Miltenyi-Biotec) spanning the entire spike (RBD/S1/S2) protein (S), nucleocapsid phosphoprotein (NP) and membrane glycoprotein (M). Additional tubes containing phytohaemagglutinin-L (Sigma) or nothing were run alongside as positive and negative controls respectively. Whole blood samples were incubated at 37°C for 20-24 hours before harvesting plasma from each sample to quantify IFN-g by ELISA (BioLegend). SARS-CoV-2-specific T-cell responses were identified as positive using previously defined criteria for differentiating naïve controls from prior COVID-19 vaccinated and/or infected individuals (sensitivity 96.0% and specificity 84.4%) [5].

### Statistical analysis

Humoral and T-cell response rates to COVID-19 vaccine 3 are expressed using descriptive statistics. Fisher’s Exact test and a two-sample t-test were used to explore the relationship between prior COVID-19 infection and immune response to COVID-19 vaccine 3. Statistical analysis was performed in Stata v16 (Stata corp ltd).

## Results

Of 188 potential participants invited, 81 participated. 79 participants provided a dried blood spot sample, of whom 38 also provided a whole blood sample; 2 provided only whole blood. Fifty-eight (72%) were women, mean age 45.8 years, 55 were receiving ocrelizumab and 15 fingolimod, 9 were on other immunosuppressants (in some cases for co-morbidity rather than MS) and 2 received no DMT (Table 1). Mean interval from vaccine 3 to blood draw was 5.9 (SD 1.4) weeks.

**Table 1:**
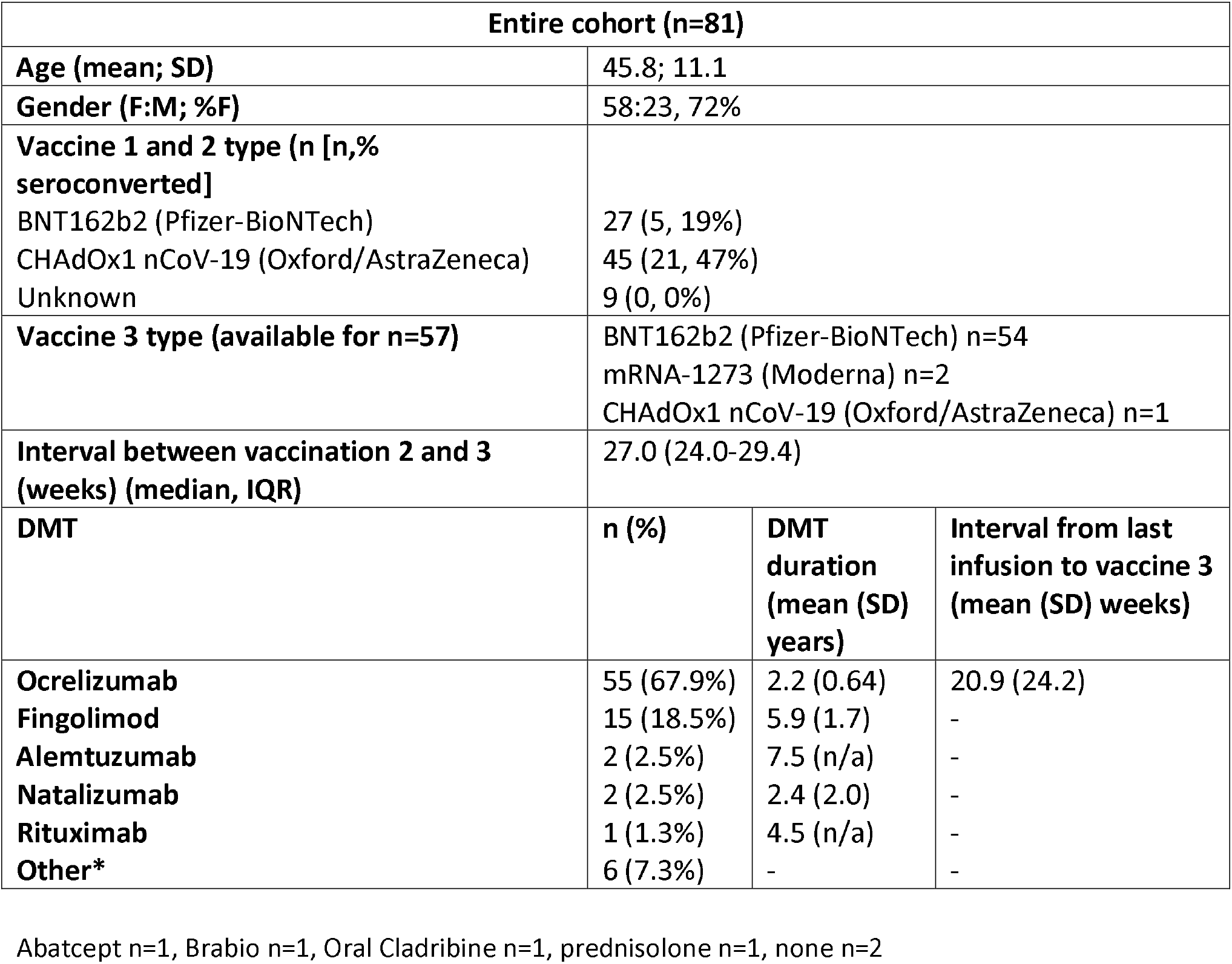
Clinical and demographic features of the study population

Anti-spike IgG data from dried blood spot samples (EuroImmun assay) demonstrated that 26 of 79 (33%) participants had seroconverted post-COVID-19 vaccine 3. Eight of 52 (15%) people receiving ocrelizumab seroconverted, and 7 of 15 (47%) people taking fingolimod. Six people had borderline results: 3 on ocrelizumab and 3 on fingolimod. People who received the CHAdOx1 nCoV-19 (Oxford/AstraZeneca) vaccine for their initial (first and second) vaccination were more likely to seroconvert that those who received the BNT162b2 (Pfizer-BioNTech) for their initial vaccine (odds ratio, OR 3.85, 95% confidence interval 1.24-11.97, p=0.02) (Table 1).

Of the 38 participants who had paired whole blood samples (Bio-rad assay), concordance for Euroimmun seropositivity was 100%, but an additional 9 (24%) people seronegative on DBS analysis were found to be anti-spike positive (Figure 1), giving a Kappa coefficient for agreement of 0.76. The assays appeared to have good correlation (r^2^=0.89), which persisted when only samples with a positive result on at least one assay were included (r^2^=0.86) (supplementary data 1).

**Figure 1:**
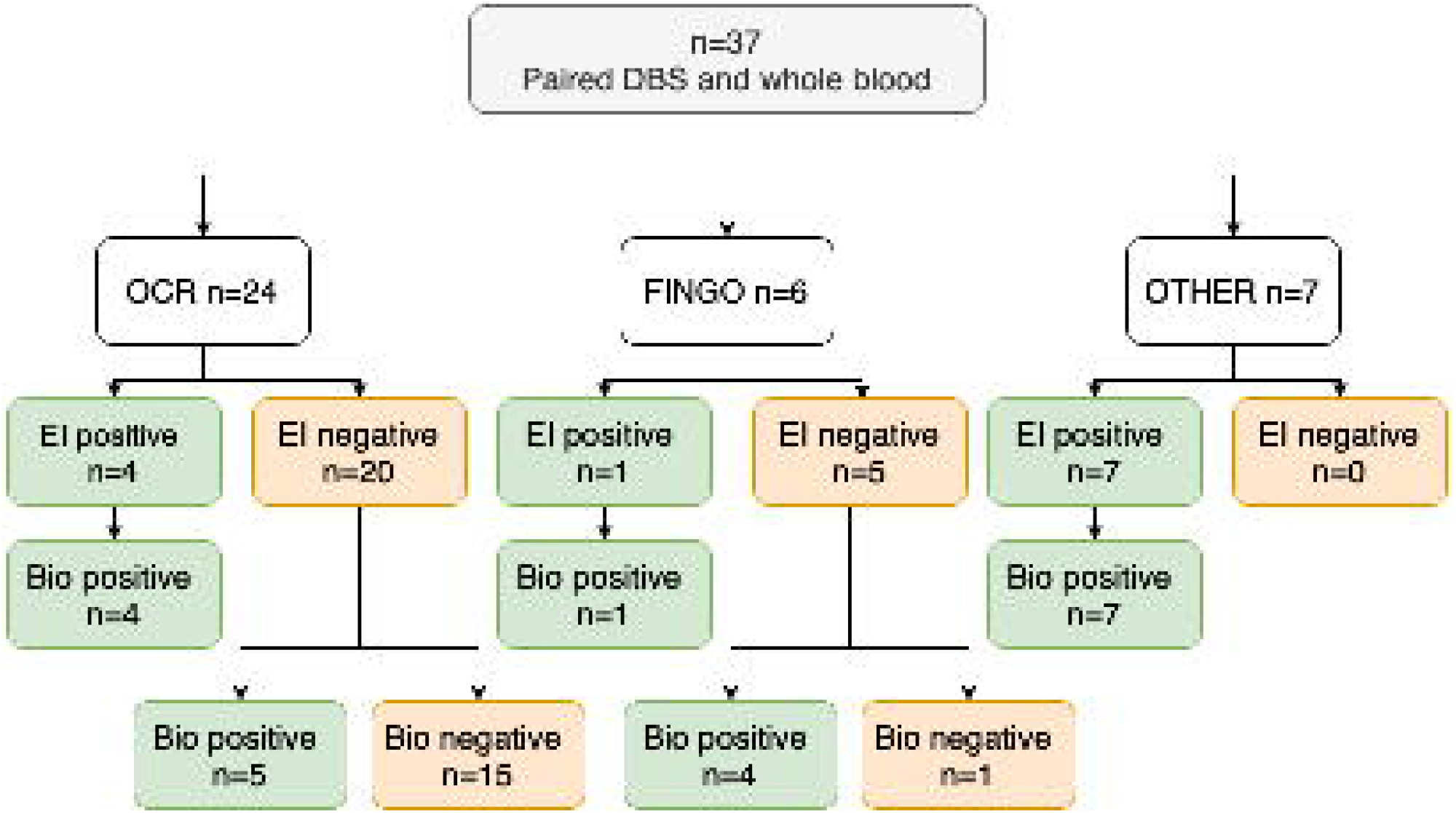
Flow diagram illustrating seropositive / seronegative status to two anti-spike IgG assays. Bio: Bio-Rad anti-spike IgG assay performed on whole blood, DBS: dried blood spot, EI: EuroImmun anti-spike IgG assay performed on dried blood spots, Fingo: fingolimod, OCR: ocrelizumab, OTHER: see Table 1 for range of other immune-modulating treatments.

Anti-spike T-cell responses were positive in 26/40 (65%) participants, including 24/28 (86%) on ocrelizumab and 0/6 (0%) on fingolimod (Figure 2). Overall, 31/40 (78%) demonstrated either humoral or cellular immune response post-COVID-19 vaccine 3; this rose to 35/40 (88%) when including IgG results from whole blood.

**Figure 2:**
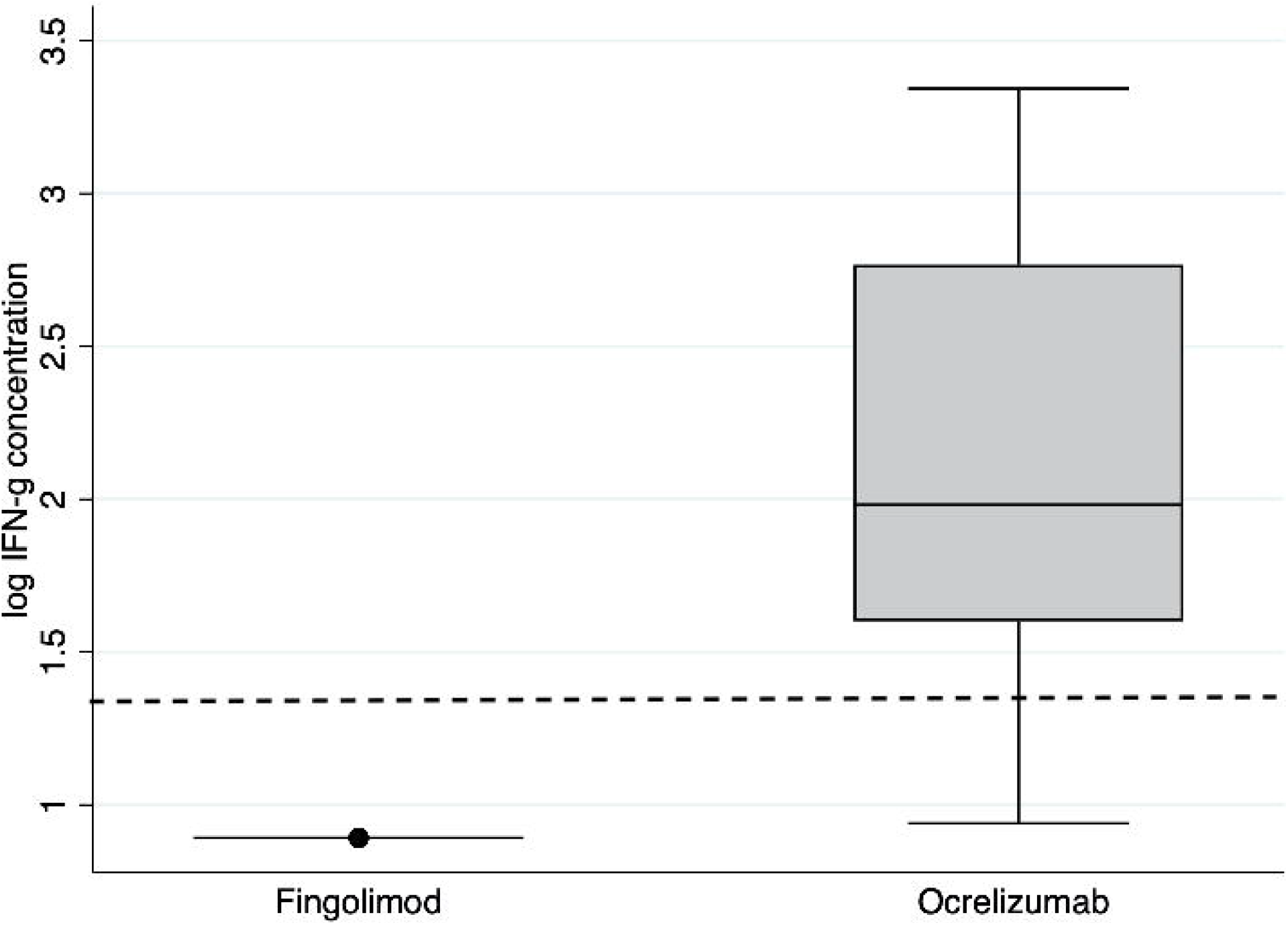
Box and whisker plots of quantitative anti-spike T-cell responses (IFN-g concentration in plasma following stimulation) according to disease modifying therapy (fingolimod [n=6] vs ocrelizumab [n=28]). The box represents the interquartile range intersected by the median; whiskers the range. The dotted line represents the positive/negative cut off value.

Laboratory evidence of COVID-19 infection (either antibody or T-cell response to nucleocapsid) was present in 14/40 (35%), of whom 2 had prior laboratory-confirmed COVID-19. There was no association between presence or absence of laboratory evidence of prior COVID-19 infection and either T-cell response or anti-spike seroconversion following COVID-19 vaccine 3 as measured by Fishers exact test. However, evidence of prior infection appeared to be associated with a greater T-cell response (p<0.0001, 2 sample t-test) (Figure 3a). Following the exclusion of 2 outliers with very low IgG responses, there was no significant difference in quantitative IgG response between those with and without evidence of prior infection (p=0.33) (Figure 3b). Six of 81 (7.5%) participants experienced PCR-confirmed COVID-19 infection subsequent to vaccine 3 and blood sampling. Four of these six had either T-cell or antibody response to SARS-CoV-2 spike when tested prior to developing COVID; all recovered without antivirals or hospital admission.

**Figure 3:**
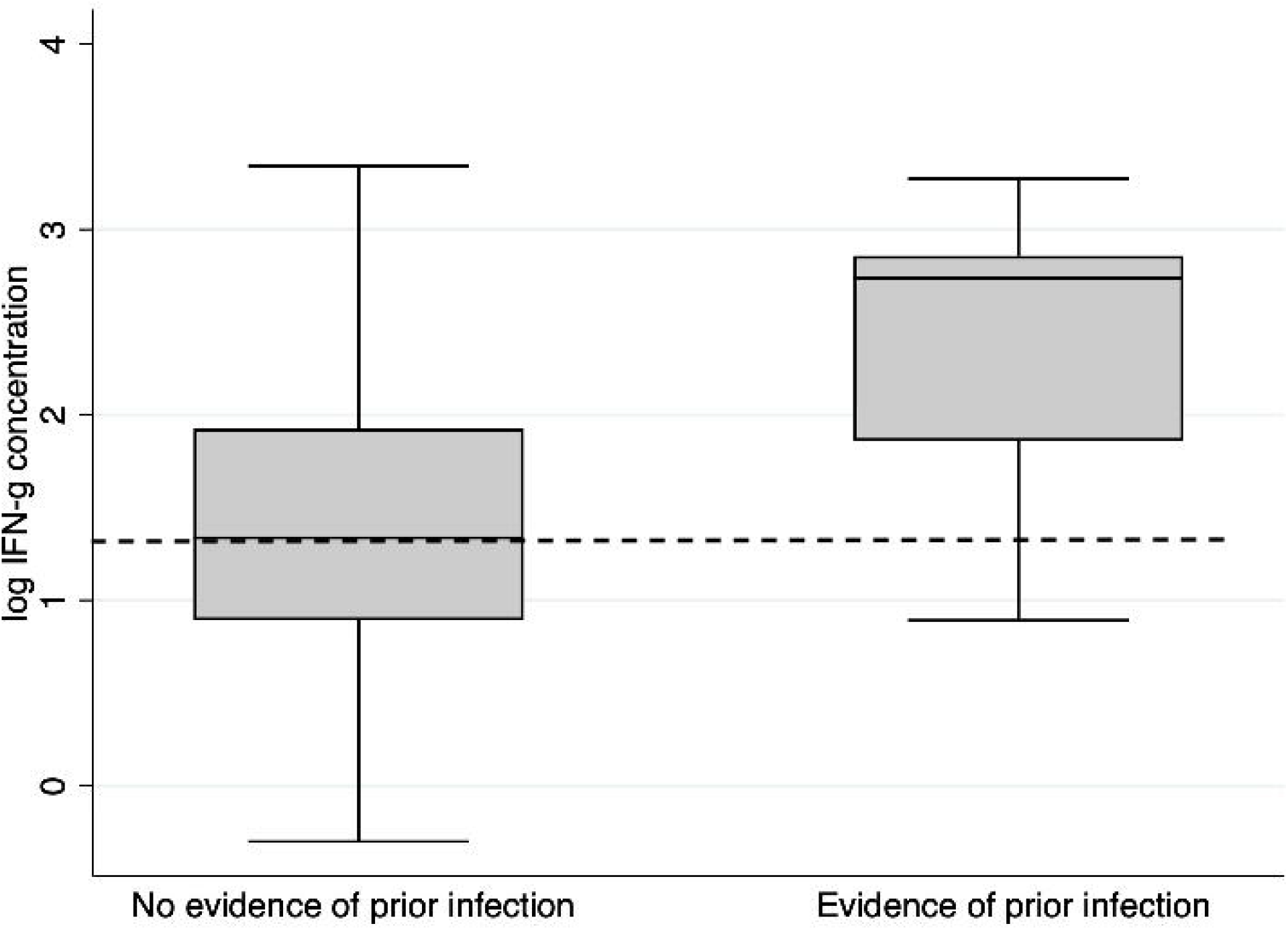
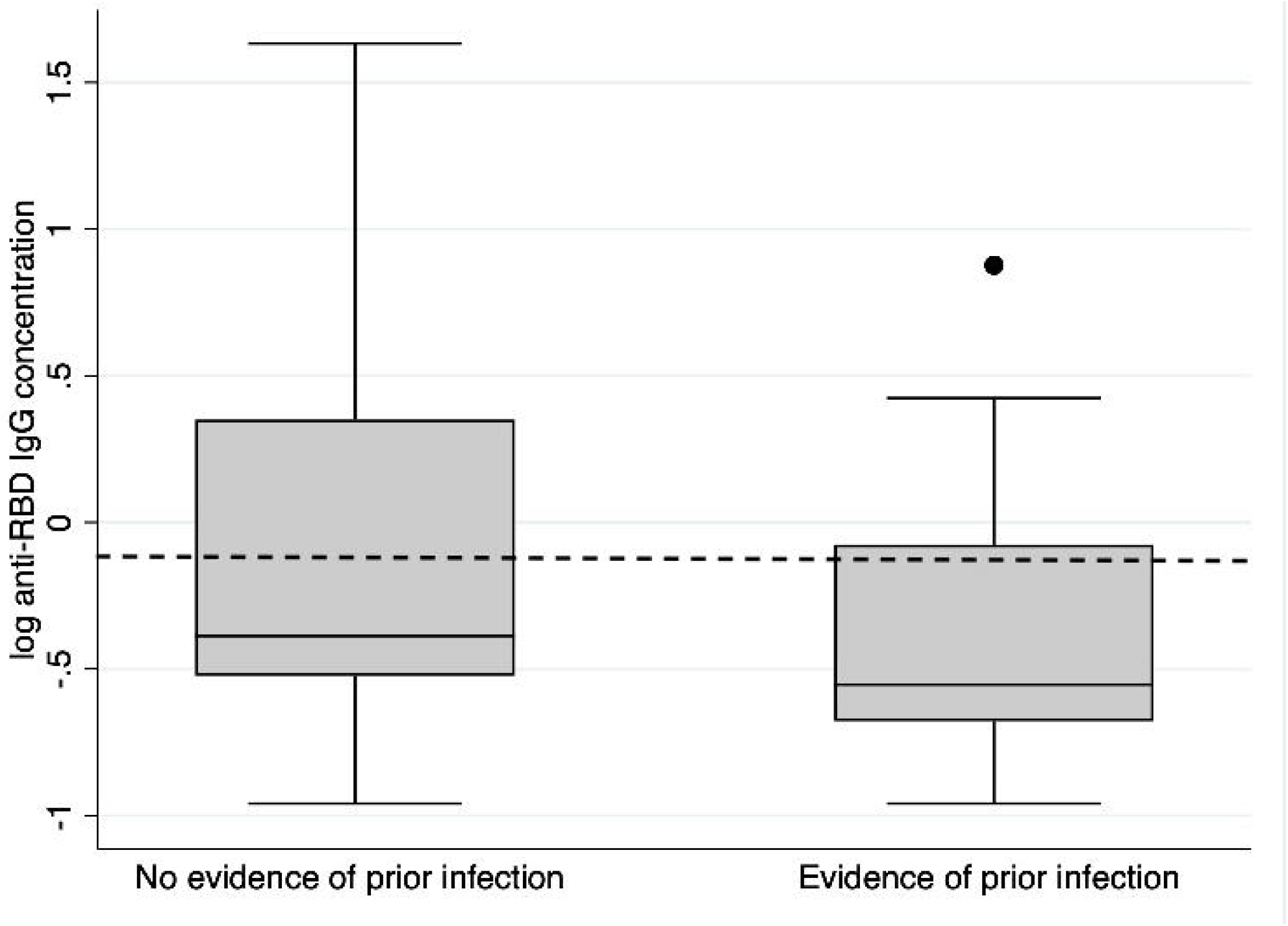
Box and whisker plots of (a) quantitative anti-spike T cell responses (IFN-g concentration in plasma following stimulation) and (b) quantitative anti-spike IgG titres measured using the EuroImmun ELISA on dried blood spots according to evidence of prior COVID-19 infection (defined by either T-cell or antibody responses directed against the nucleocapsid antigen). N=26 samples had no evidence of prior infection, and n=12 had evidence of prior infection. The box represents the interquartile range intersected by the median; whiskers the range. The dotted line represents the positive/negative cut off value.

## Discussion

We show that a third of pwMS who were seronegative after initial COVID-19 vaccination seroconverted after booster (3rd) vaccination, and between 8 and 9 out of 10 had a measurable immune response to COVID-19, depending on the IgG assay used. These findings support the use of booster vaccinations in this group. Many pwMS exposed to disease modifying therapies (DMTs) have an attenuated response to initial COVID-19 vaccination. This has been most profound when measuring humoral immunity in those exposed to anti-CD20s, and fits with data showing an increased vulnerability to COVID-19 amongst this group [6-8].

Our findings are aligned with the mechanistic action of the MS DMTs. Seroconversion after COVID-19 vaccine 3 occurred in a high proportion of those on fingolimod, and around 1 in 6 of those on anti-CD20 therapy, in keeping with another recent study of humoral response [9]. Conversely, T-cell responses were not demonstrated in any of the 6 people we studied on fingolimod but were present in the majority of those on ocrelizumab. Other studies have demonstrated robust or even augmented T-cell responses to either COVID-19 vaccines 1&2 [10-12], and to COVID-19 infection in pwMS on antiCD20 therapy [13]. In our sample of pwMS, three had laboratory confirmed COVID-19 after vaccine 3 but all cases were mild. It is possible that the T-cell responses seen following booster vaccination were initially generated following the initial vaccine course, however the improvement in the proportion who mount a serological response to vaccination provides a rationale for people with MS to take up booster vaccinations.

We found that the proportion of people who seroconvert following vaccine 3 depends to some degree on the assay used. This highlights the importance of assay selection, and the consideration of borderline values when interpreting results. The implications for clinical immunity in those with borderline IgG titres, i.e. those people who test positive on one assay and negative on another, remains to be determined [9]. Furthermore, the finding that none of the participants taking fingolimod generated a detectable T-cell response requires further thought. It may be that this response is truly absent, or that there are too few circulating T-cells to allow detectable responses. Further explanations could be that the relevant T-cells are trapped in secondary lymphoid organs due to the mode of action of fingolimod, or that circulating fingolimod in whole blood samples blocks in vitro responses.

The clinical correlates of antibody and T-cell responses to COVID-19 remain uncertain but exposure to antigen provoking measurable antibody and T-cell responses is almost certainly associated with milder subsequent disease [14]. Virus-specific CD4+ and CD8+ T cell responses produce effector cytokines and exert cytotoxic activity. Antibodies can directly neutralise virus and reduce viral load, or effect functions including antibody-dependent cellular cytotoxicity and complement deposition. Recent studies have demonstrated that newer SARS-CoV-2 variants appear less susceptible to the neutralising activity of COVID-19 vaccine-elicited antibodies [15-17]. On the other hand, SARS-CoV-2-specific T-cell epitopes appear to be conserved among emerging variants [18], suggesting that T-cell responses may be particularly important in attenuating current COVID-19 severity.

Higher incidence and/or worse outcomes in the context of in vivo COVID-19 infection in people receiving anti-CD20, [5,6] and fingolimod,[7] have been observed in some large population-based studies performed prior to the widespread availability of booster vaccines. These observations require further study to fully understand the clinical implications of booster vaccination and their underlying mechanisms.

## Conclusion

The third dose of COVID-19 vaccination appears to provide additional benefit to people with MS who have failed to respond to the initial vaccine course. All people with MS should therefore be encouraged to follow vaccination schedules in order to obtain maximal possible protection. T-cell and antibody testing of pwMS on certain DMTs may allow more individualised counselling on infection-risk. However, the clinical correlates and durability of these immune responses will require further longitudinal studies.

## Data Availability

Anonymised data are available upon reasonable request to the authors

---

**Supplementary figure 1:** Correlation between IgG titres as measured by EuroImmun ELISA and using the Bio-Plex Pro Human SARS-CoV-2 – all samples

**Supplementary figure 2:** Correlation between IgG titres as measured by EuroImmun ELISA and using the Bio-Plex Pro Human SARS-CoV-2 – samples testing positive on Bio-Plex Pro Human SARS-CoV-2 only

## Acknowledgements

The T cell work was funded by BMA Foundation for Medical Research (Vera Down award) to RD and ECT. The authors would like to acknowledge the help of Aliye Nazli Asardag, Sita Navin Shah, Swee Vickie Nixon, Ray Wynford-Thomas, Marija Cauchi, Catherine McConnell, Cynthia Butcher, Zin Min Htet, Miranda Maria Piana Parra, Joseph Bishop and Raees Samli in the running of the study.

## Additional study support was provided as follows

**UHW:** Support for equipment and consumables was provided by Cardiff and Vale UHB and Cardiff University. Salary for Samantha Loveless was partly provided by the BRAIN Unit Infrastructure Award (Grant no: UA05). The BRAIN Unit is funded by Welsh Government through Health and Care Research Wales.

**QMUL:** Work performed as part of this study at QMUL is funded by Merck Serono Ltd., Feltham, UK, an affiliate of Merck KGaA and Biogen UK. There was additional support from the MS community via crowdfunding. This work was performed within the PNU, which is funded by Barts Charity.

## Author contributions

ECT, RD, SJ, NPR, NV, GG, AG, KS, MS, DB, MW and SM contributed to the conception and design of the study; NV, AR, VA, RC, NE, KG, AG, KEH, AH, GI, MJ, AK, SJM, MS, and SL contributed to the acquisition and analysis of data; ECT, NV, RD, MS and AG contributed to drafting the text or preparing the figures.

