## Supplementary figures and images for "Response to COVID-19 booster vaccinations in seronegative people with MS"

### Supplementary figure 1

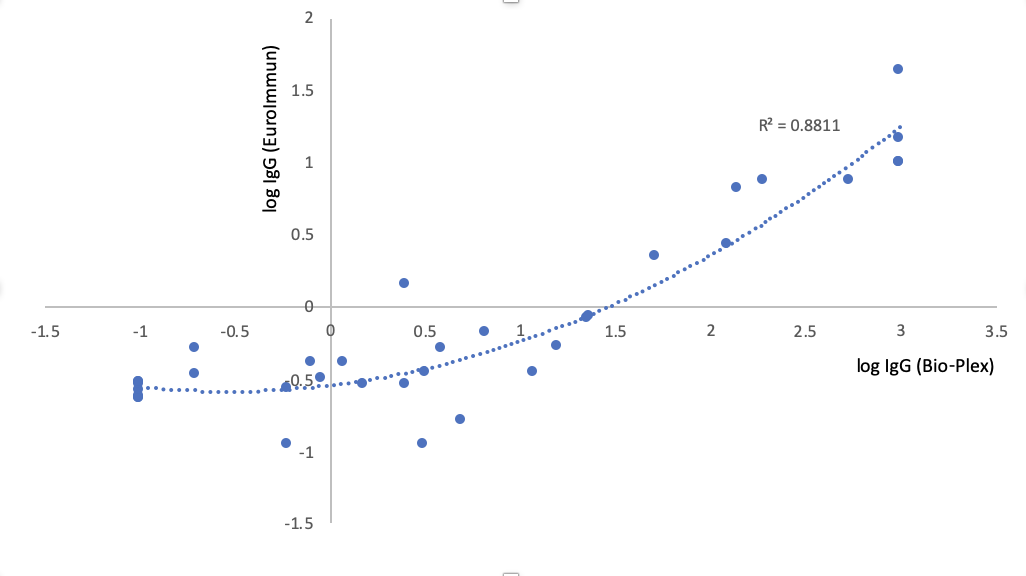

### Supplementary figure 2

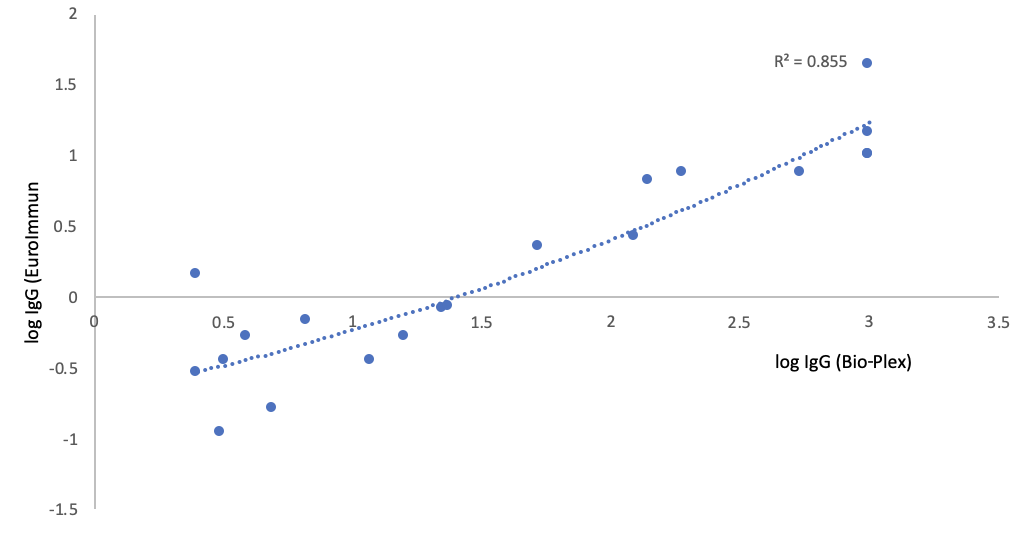
